# The McGill Approach to Core Stabilization in the Treatment of Chronic Low Back Pain: A Review

**DOI:** 10.1101/2022.01.21.22269311

**Authors:** Erica Laurin, Amir Minerbi, LCol Markus Besemann, Captain Isabel Courchesne, Gaurav Gupta

## Abstract

**Introduction:** Low back pain (LBP) is a major cause of disability and is progressively becoming worse on a global scale. [1,2] The prevention and rehabilitation of LBP lacks clarity in part due to the heterogeneity of the exercise programs prescribed to treat low back pain. Some authors have proposed stabilizing exercises for lower back pain which exert minimal loads on the spine. [3,4,5] Despite a multitude of existing exercise therapies, McGill has introduced three exercises for rehabilitating lower back pain, termed the McGill Big Three (MGB3). [6,7,8,9,10] These include the curl-up, side plank and bird-dog. The purpose of this review is to investigate the clinical outcomes from prescribing the MGB3 to individuals with chronic LBP.

**Methods:** Inclusion criteria were randomized control trials that involved an intervention with MGB3 core stabilization exercises for patients with chronic low back pain. The research included articles published during any period in full English text. Studies were critically reviewed by two authors EL and GG independently and collaboratively.

**Results:** In total, four randomized control trials were included in this review. Multiple cohorts, with varying age, demographics and occupation were studied. Outcomes studied included various pain scores, patient reported functional and performance measures.

**Discussion:** Controlled clinical trials employing this method in low back pain treatment showed low quality data with mixed statistical significance, and little to no clinical significance irrespective of the measure used or even when compared to baseline. Limitations of these trials are detailed herein.

**Conclusion:** Currently there is limited data supporting the clinical benefit of the McGill approach for the treatment of low back pain based on the available randomized clinical trials. More study is required prior to widespread adoption into clinical practice.

## Introduction

Chronic low back pain (LBP), generally referred to as pain or stiffness in the lumbar region of the spine lasting a minimum of 12 weeks, is among the leading causes of disability globally. [11,12,13,14,15, 16,17] LBP poses an increasing medical and economic burden, attributable primarily to treatments costs and decreased work productivity. [1,2] The point prevalence and lifetime prevalence of LBP among the Canadian population is 28.4% and 84.1%, respectively. [18,19] The prevalence of chronic back issues within the Canadian Armed Forces (CAF) is 16%, and LBP specifically accounts for 12% of medical releases each year. [20, 21]

Mechanical LBP is hypothesized to occur from anatomical structures such as the muscles, ligaments, bones, joints, spinal nerve roots, intervertebral discs and abdominal organs. [22,23,24] Associated precursors for mechanical LBP can include traumatic injury, structural abnormalities, pregnancy, infection, inflammatory disorders, degenerative conditions, disc herniation, radiculopathy and spinal stenosis. [25,26] Risk factors for LBP may include specific types of repetitive spinal motion, low socioeconomic status, medical and psychiatric comorbidities, and inadequate coping mechanisms. [15,27, 28,29] There is also emerging evidence indicating LBP is not only a regional issue given the studies supporting altered cerebral functional gray and white matter in patients, although the directionality and temporality have yet to be established. [30] Treatments include but are not limited to pharmaceuticals, exercise therapy, manual therapies, injections and surgery. [15,25,31,32]

Possibly owing to the various underlying causes of LBP, heterogeneity of evaluation tools and measures of success, some authors argue it is nearly impossible to formulate standardized exercise interventions to treat chronic LBP. [5,6,25,33,34]. However, within prescribed exercise therapies for LBP, core stabilization exercises have gained increased attention over the recent decade. [5,35,36,37] This type of exercise therapy has evolved into a mainstay for treating chronic LBP with focus on core stabilization and muscle coordination. [3,38,39,40,41,42,43]

Spinal stability is theorized to be an integration of active muscle contraction, passive ligament support and neural control. [39,40,42] Active muscle control can be further classified into local (i.e., segmental stability) and global stabilizers and dynamic control. [39,42,43]. Core stabilizing exercises train patterns of muscle activity and posture without producing excessive loading of the spine. [44,45] Stability is determined by various motor patterns that differ in aspects such as demand, load and speed. [45]

Therefore, it is of great interest to validate the most clinically effective rehabilitation program for individuals with LBP. Various approaches have been studied including generalized flexibility, variable load anterior/posterior muscle training, along with the low to no load lumbo-pelvic region based on the global muscle stabilization described by McGill. [40,41,42,43] Based on numerous biomechanical and clinical studies, McGill has proposed stabilizing exercises which apply minimal loads on the spine to reduce and prevent LBP. [3] This includes tailored programs built around three exercises termed the “McGill Big Three” (MGB3) which include the curl-up, side plank and bird-dog (please see Figure 1 for examples of these exercises). [10]

**Figure 1:** The McGill Big “3” Core Stabilization exercises. Figure 1.1. Curl-up Pls see https://www.healthaya.com/publications Figure 1.2. Side-plank Pls see https://www.healthaya.com/publications Figure 1.3. Bird-dog Pls see https://www.healthaya.com/publications

In order to further delineate the clinical impact of the McGill approach in the treatment of chronic low back pain, we undertook a review of the existing literature. To our knowledge this systematic evaluation has not been completed thus far.

## Methods

A review of the literature was completed using the following search terms: McGill stabilization exercises, McGill Big Three, MGB3, low back pain (LBP), Stuart McGill, chronic low back pain (LBP), core stabilizing exercises. The PubMed, Ovid Medline, Embase and Google Scholar electronic databases were used. References of existing articles were inspected to identify additional relevant studies. Inclusion criteria were randomized control trials that involved an intervention with MGB3 core stabilization exercises for patients with chronic low back pain. The research included articles published during any period in full English text. Studies were critically reviewed by two authors EL and GG independently and collaboratively.

For the analysis, between group differences were calculated by using the absolute differences for various performance measures and patient reported pain and functional outcomes. For instance, if mean pain scores in the control group decreased from 8/10 to 4/10, this was considered to be a 40 percent improvement (not 50%), which was subsequently subtracted from the corresponding measure in the treatment group. Where range of motion was used, the total possible score was calculated using a maximum of 20 degrees of extension and 40 degrees of flexion, for a total range of motion of degrees [46]. Percentage improvement was calculated against degrees improvement divided by 60 degrees. One study provided mean scores using several graphs. [48] Percentage improvement for this study was therefore calculated from estimated scores. Finally, where multiple performance measures were used, the measure with a total possible maximum score was used. [3,43,47,48] A positive score represents an effect favoring the McGill approach for a given study. A trial reporting performance measures using covariate analysis was excluded from the analysis as the corresponding author was unable to provide mean scores. [3]

## Results

In total, four randomized control studies satisfied the inclusion criteria. [3,43,47,48] 170 participants were recruited from the four studies with 23 lost to follow up. Participants ranged from 20 to 60 years of age. Plausible confounding variables including gender, age, weight and height were recorded in all studies, and the assessors were blinded to treatment allocation with the exception of Ghorbanpour et al. 2018. Ammar et al. 2011 recruited only postnatal female participants and Chan et al. 2020 recruited male military participants only.

Chan et al. 2020 provided 5 weeks of pre-trial passive pain treatment with heat and transcutaneous electrical stimulation (TENS). Ammar 2012 provided heat to all participants, while Ammar et. al 2011 provided heat to the control group only. No studies used non-treatment control groups, and protocols for the prescribed exercises varied with respect to type of exercises, duration, and intensity of exercises for both groups. Furthermore, the McGill stabilization exercises were not uniform throughout each study as Ghorbanpour et. al 2012 and Chan et al. 2020 focused only on the MGB3, while Ammar 2012 and Ammar et. al 2011 employed McGill stabilization exercises beyond the MGB3. [3,47,48,43] Treatment outcomes were measured at 4 or 6 weeks depending on the trial, and no longer-term measures were assessed in any trial. [3,43,47,48]

Two studies, Ammar et al. 2011 and Ammar 2012 monitored compliance using patient reported logs and were the only studies that showed statistical differences for outcomes between groups. Ammar et. al 2011 insufficiently differentiated groups, and mean scores and standard deviations for outcomes were identical for one group suggesting a possibility of error in the results. Ghorbanpour et al 2018 appear to have errors when reporting mean difference which were adjusted for in our analysis. Chan et al. 2020 was the only study conducted with a therapist present for all sessions. The same study provided a power analysis to justify recruitment and accounted for dropouts in their analysis. When looking at the relative improvements between groups, pain, function and performance scores saw an improvement of <10% across the studies, with the exception of Chan et. al which saw an improvement of 15% for pain scores. Table 1 describes the specifics of the studies while Figure 2 details the comparative differences between studies on select outcome measures.

**Table 1:** General Summary of Studies Included in Review.

| Study | Population (n) | Intervention for Control Groups | Patient/ Inclusion Criteria | Exclusion Criteria | Time of Intervention | Outcome Measurements | Results |
| --- | --- | --- | --- | --- | --- | --- | --- |
| Ammar (2012) | 60 (initially 67 – 7 dropouts)<br>G1 = Traditional (n=30)<br>G2 = McGill (n=30) | Conventional exercises (stretching and strengthening exercises for trunk and lower limbs)<br>Mixed home program and in-clinic program | M/F<br>CNSLBP<br>Ages 29-60<br>M/F<br>G1: 11/19<br>G2: 13/17 | History of previous lumbar surgery, spinal stenosis, systemic disease, spondylolisthesis, neurological dysfunction, injection therapy, carcinoma or pregnancy | T0 = baseline<br>T1 = 6 weeks | Fifty-foot speed walk, a fifty-foot fast speed walk and distance walked in five minutes<br>Compliance measured by logs | Physical Function:<br>G2 > G1<br>statistically significant between groups |
| Ammar et. al (2011) | 34 (initially 39 – 5 dropouts)<br>G1 = Traditional (n=17)<br>G2 = McGill (n=17) | Heat, Stretching and strengthening exercises for trunk and lower limbs<br>Trunk flexion, extension, rotation, lateral flexion, Hip extension, stretching piriformis, hip abductor and extensor<br>Mixed home program and in-clinic program | F<br>18 years and older with postnatal LBP<br>Ages 21-38 years old | History of previous lumbar surgery, spinal stenosis, spondylolisthesis, neurological dysfunction, radiculopathy, systemic disease, carcinoma, injection therapy, or a reluctance to participate in the study | T0 = baseline<br>T1 = 4 weeks | Numerical pain rating scale<br>Oswestry disability questionnaire<br>Compliance measured by logs | Pain:<br>G2 > G1<br>Disability:<br>G2 > G1<br>statistically significant between groups |
| Ghorbanpour (2018) | 30 (initially 34 – 4 dropouts)<br>G1 = PT (n=17)<br>G2 = McGill (n=17) | Conventional PT single and double knee to chest prone lying with pillow with one leg sliding, cycling in supine and bridging exercises<br>Home based program<br>Not blinded | M/F<br>CNSLBP (> 6 months) without radiating pain to the leg, and no physiotherapy treatment<br>Age 20-40<br>M/F<br>G1: 7/8<br>G2: 7/8 | History of pelvic, spine, upper or lower extremities surgery, cardiovascular diseases, hamstring and quadratus lumborum muscles shortening, pain or disability in upper and lower extremities, frequent neurological deficits, and professional athletes | T0 = baseline<br>T1 = 6 weeks | Visual analog scale (VAS)<br>Persian version of Quebec Low Back Pain Disability Scale Questionnaire<br>Baseline Bubble Inclinometer | Pain:<br>G2 > G1<br>Disability:<br>G2 > G1<br>Flexion:<br>G1 > G2<br>Extension:<br>G2 > G1<br>Within group statistical differences but not between group |
| Chan et. al (2020) | 30 (7 dropouts but all participants used in analysis)<br>G1 = DMST (n=15)<br>G2 = McGill (n=15) | DMST<br>Standard pain management therapy (heat treatment using hydro collator and transcutaneous electrical nerve stimulation)<br>Supervised in-clinic program | M<br>Military personnel aged 18-42 years old with CNSLBP (>12 weeks)<br>Ages 24-42 | Specific neurological disorder, history of lumbar spine or abdominal surgery, regular painkiller consumption within 3 months duration, inability to fulfill follow-up appointments or comply with the rehabilitation program | T0 = baseline<br>T1 = 3 weeks<br>T2 = 6 weeks | Numeric pain rating scale (NRPS)<br>Malay version of Roland Morris Disability Questionnaire (RMDQ)<br>Time-based static hold test<br>Sahrmann 5-level core stability test<br>Y-Balance test | Pain:<br>G1 = G2<br>Disability:<br>G1= G2<br>Power analysis done<br>No statistically significant between groups |

**Abbreviations:** G; Group, T; Time of evaluation; Traditional; Traditional LBP Exercises; Performance; Performance Based Measures; CNSLBP; Chronic Non-Specific low back pain; M; Male; F; Female; PT; Physiotherapy; DMST; Dynamic muscular stabilization techniques

**Figure 2:**
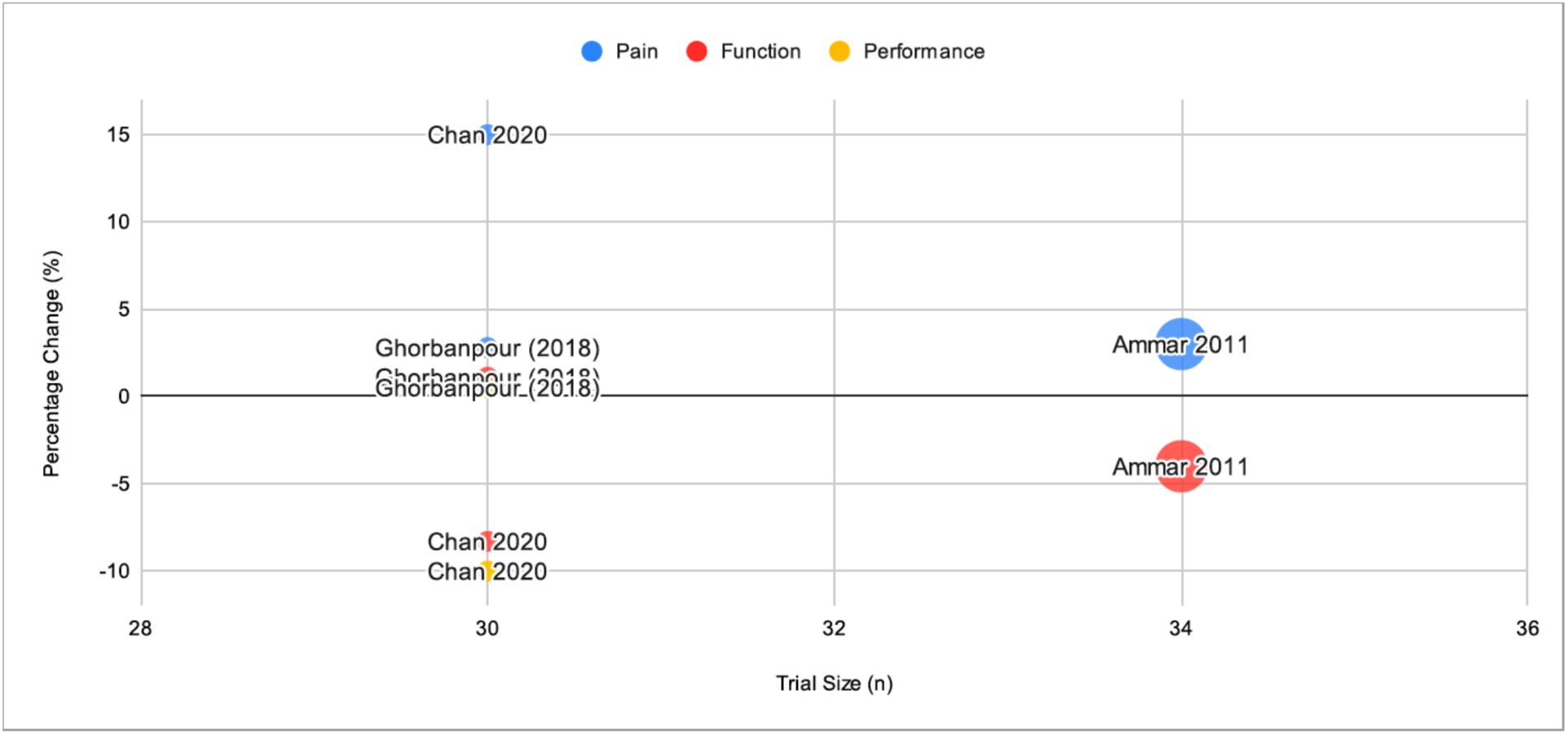
Bubble Graph Comparing Differences between Studies on Select Outcome Measures. Note: Bubble size also represents total trial size

## Discussion

Core stability training continues to be widely employed in therapy of LBP and injury prevention. The goal of this training is to stabilize the lumbopelvic region in a neutral state when performing spine-loading tasks by re-educating postural core muscles such as the transverse abdominis, multifidi, and pelvic floor muscles. It is hypothesized that this will provide segmental stability and control of the lumbar spinal segments during activity. [49, 50, 51]

There have been multiple studies looking at various types of core strengthening programs on pain and disability with mixed results. [47] As described, the McGill approach is one type of core strengthening program that incorporates the abdominal bracing strategy to recruit high trunk muscle activity with low spinal loads on the spine. [51] This review of the available randomized controlled clinical trials employing this method in low back pain treatment showed low quality data with mixed statistical significance, and little to no clinical significance irrespective of the measure used or even when compared to baseline. Limitations of these trials are detailed in Table 2. Practical limitations of the review include the exclusion of background biomechanical and cohort studies of the McGill approach, as well as the inclusion of other core strengthening approaches. This can certainly be considered in the future given the limited quality of the data available. Further investigation could consider 1) further comparing McGill or other stabilization programs to other conventional physical therapies 2) looking at these treatments alongside other types of treatments of low back pain 3) longer duration studies with carefully selected patients and robust treatment protocols and 4) impact on resource healthcare utilization and care seeking.

**Table 2:** Limitations of Trials Included in Review of McGill Approach for Low Back Pain Treatment.

| Limitation | Summary |
| --- | --- |
| Generalizability | • Pregnancy involves hormonal changes and a wide range of postpartum timings where back pain is expected to resolve<br>• Limited to younger age range therefore unclear if older patients can do the exercises as described,<br>• Unclear of utility in patients with other pain issues or comorbidities<br>• Cost effectiveness and broad clinical application unclear<br>• Only single male dominant military cohort<br>• Impact of home programs versus in person care unclear |
| Clinical significance | • Studies did not meet the generally accepted minimally clinically important difference for various measures employed |
| Heterogeneous treatment protocols | • Variable exercises, intensity, duration and progression |
| Program Tailoring and Progression | • Generalized programs prescribed<br>• How and why exercises were progressed not clear and may be a confounding factor |
| Compliance | • Only measured by self-report if at all |
| Short term outcomes/application | • Longer term outcomes past 6 weeks unclear<br>• Literature suggests better outcomes with longer duration treatment [52] |
| Heterogeneous assessment tools | • Various measures of pain, function and performance used with unclear clinical validation in some cases |
| Heterogeneous controls groups | • No studies used non-treatment controls, therefore the natural and variable course of low back pain could not be included |
| Co-adjuvant therapy | • Unclear how this was controlled for in studies |
| Small studies sizes and number | • Generally no power analysis stated |

## Conclusion

Currently there is limited data supporting the clinical benefit of the McGill approach for the treatment of low back pain based on the available randomized clinical trials. Further studies are required to verify the treatment efficacy prior to widespread adoption into clinical practice.

## Disclaimer

The views expressed herein are those of the author and may not reflect those of the Canadian Armed Forces, Canadian Forces Health Services, the Department of National Defense or the Canadian Government.

## Data Availability

All data produced in the present work are contained in the manuscript

## Acknowledgements

We would like to thank Major Pauline Godsell and Captain Nicole Mahoney for their feedback and support on this project.

